# Moderate Static Magnetic Fields Prevent Estrogen Deficiency-Induced Bone Loss: Evidence from Ovariectomized Mouse Model and Small Sample Size Randomized Controlled Clinical Trial

**DOI:** 10.1101/2024.11.08.24316968

**Authors:** Shenghang Wang, Jiancheng Yang, Yunpeng Wei, Chao Cai, Shuai Chen, Youde Wu, Xiao Li, Lilei Sun, Xianglin Li, Ming Gong, Jianhua Zhou, Yawei Hu, Wang Zhang, Zengfeng Guo, Jiacheng Liao, Chunling Luo, Xiaosu Bai, Xinle luo, Liming Duan, Ting Huyan, Zhouqi Yang, Min Wei, Yanwen Fang, Hao Zhang, Peng Shang

**Affiliations:** Department of Spine Surgery, People’s Hospital of Longhua, Shenzhen, China; Research & Development Institute of Northwestern Polytechnical University in Shenzhen, Shenzhen, China; Department of Osteoporosis, Honghui Hospital, Xi’an Jiaotong University, Xi’an, China; Shenzhen University of Advanced Technology, Shenzhen, China; School of Life Sciences, Northwestern Polytechnical University, Xi’an, China; Minle Community Health Service Center, People’s Hospital of Longhua, Shenzhen, China; Department of Endocrinology, People’s Hospital of Longhua, Shenzhen, China; An Shi Rui Information Technology Co., Ltd, Shenzhen, China; Heye Health Technology Co., Ltd, Huzhou, China

**Author notes:** Corresponding author: Peng Shang, Research & Development Institute of Northwestern Polytechnical University in Shenzhen, No. 45, Gaoxin South 9th Road, Nanshan District, Shenzhen, China., Hao Zhang, Department of Spine Surgery, People’s Hospital of Longhua, Affiliated Hospital of Southern Medical University, No.38 Jinglong Construction Road, Shenzhen, China. These authors contributed equally to this work (Shenghang Wang, Jiancheng Yang).

**Keywords:** Moderate static magnetic field, Ovariectomized mice, Postmenopausal osteoporosis, Bone turnover

## Abstract

**Background:** Postmenopausal osteoporosis (PMOP) is the most common type of osteoporosis. Numerous studies have shown that static magnetic fields (SMFs) can inhibit bone loss by regulating bone remodeling. However, there are currently no clinical studies on the treatment of osteoporosis with SMFs. This study aims to investigate the clinical therapeutic effects of moderate static magnetic fields (MMFs) on PMOP.

**Methods:** In this paper, we constructed MMF device using neodymium-iron-boron (NdFeB) materials. At the animal level, the effect of MMF exposure for 8 weeks on estrogen deficiency-induced bone loss was investigated by evaluating bone microstructure, mechanical properties, and bone conversion using ovariectomized (OVX) mice. Clinically, a single-blind randomized controlled study in patients with PMOP was designed. PMOP patients aged 55-70 years were recruited and randomized into the control and MMF treatment groups. Clinical assessments of bone mineral density (BMD), bone turnover markers (BTMs) and VAS scores were performed at baseline and day 90, respectively.

**Results:** The results showed that MMF exposure significantly improved BMD, bone mineral content (BMC), bone microarchitecture and bone strength in OVX mice. For bone turnover, MMF increased the number of osteoblasts on the bone surface of OVX mice as well as the level of serum bone formation marker P1NP, while decreasing the number of osteoclasts and the level of serum bone resorption marker β-CTX. The clinical trial’s results showed that MMF treatment had a positive effect on the improvement of BMD in the lumbar spine and increased serum P1NP levels while decreased β-CTX levels. In addition, MMF treatment decreased participants’ VAS scores for low back pain.

**Conclusions:** The results of both animal and clinical studies demonstrated that MMF treatment improved bone turnover and have a positive effect on BMD improvement, as well as alleviated low back pain in PMOP patients. This study will promote the translational research and clinical application of SMF treatment for osteoporosis.

**Trial registration:** Intervention study of moderate static magnetic field on osteoporosis and iron metabolism in postmenopausal women, ChiCTR2100048604

## Introduction

As the most common skeletal metabolic disease, osteoporosis (OP) is generally considered to be one of the factors contributing to the increased risk of fracture due to low bone mass and deterioration of bone microarchitecture[1]. Postmenopausal osteoporosis (PMOP) is caused by estrogen deficiency after women menopause, which is the most common type of age-related osteoporosis[2]. PMOP is a significant contributing factor to fractures in elderly women, and studies have indicated that the probability of fractures in women over the age of 50 is average up to 50%[3, 4]. PMOP associated with population aging becomes a major clinical and public health problem.

Currently, medication is still the mainstay of osteoporosis treatment, with commonly used drugs including the anti-resorptive drugs such as bisphosphonates, denosumab, calcitonin, and selective estrogen receptor modulators, as well as the anabolic drugs such as teriparatide, romosozumab, etc.[5–7]. Physical therapy is an indispensable adjuvant therapy for the treatment of osteoporosis[8]. Electromagnetic field is an important treatment method for orthopedic diseases, and has been used for a long history spanning several decades. Pulsed electromagnetic field (PEMF) therapy has been approved for clinical adjunctive treatment of nonunion fractures and osteoporosis[9, 10]. However, it still suffers from drawbacks such as high cost and limited portability.

Static magnetic fields (SMFs), as an easily obtainable physical factor, has been applied in the treatment research of orthopedic diseases for half a century[11–13]. Compared to other types of electromagnetic fields such as PEMF and radiofrequency electromagnetic fields (RFs), SMFs exhibit higher biosafety due to its passive nature and energy-free. Moderate static magnetic fields (MMFs) refer to SMFs with a magnetic induction intensity between 1mT and 1T, which could be acquired by permanent magnetic materials such as Samarium-Cobalt and Neodymium, which allow wide applicability at an affordable price[14, 15]. Various studies have demonstrated that MMFs could promote repair of fracture and bone defect, and inhibit bone loss, by modulating the physiological functions of osteoblast, osteoclast, and osteocyte [16, 17]. The intramedullary implant with MMFs could improve fracture healing in rabbits[18, 19]. Our preliminary research found that 0.2-0.4 T MMF whole-body exposure alleviated bone loss in OVX mice[20]. However, the clinical effectiveness of SMFs in the treatment of osteoporosis remains a mystery.

To figure out the therapeutic effectiveness and potential applications of MMFs in PMOP, we evaluated the effect of MMF on bone loss in ovariectomized (OVX) mice at the animal level; and designed a small sample size randomized controlled trial (RCT) to investigate the therapeutic efficacy of MMF on osteoporosis in postmenopausal women at the clinical level.

## Methods

### Animals and Treatments

Twenty-four adult ten-week-old female C57BL/6 mice were housed at an ambient temperature of 25°C under an artificial 12 h light/dark cycle with food and water provided. All experiment mice were randomly divided into three groups: Control group, OVX group, OVX-MMF group. After anesthesia with isoflurane, mice in OVX group and OVX-MMF group were ovariectomized, and the fat around the ovaries were excised in the mice of control group. Mice in OVX-MMF group were housed with the bottom of their cages equipped with moderate static magnetic field device. While cages of mice in OVX group were equipped with the same structural devices but without MMF.

After two weeks of the operation, the mice were treatment with MMF for 8 weeks, continuously. At the end of the eight-week treatment, the mice were euthanized by cervical dislocation under anesthesia. The blood samples and organs were collected for subsequent analyzes. All animal protocols used in this study were approved by the Lab Animal Ethics and Welfare Committee of Northwestern Polytechnical University (No.202101102).

### MMF Exposure Device for Mice

MMF exposure device for mice is composed of a plurality of N42 neodymium-iron-boron (NdFeB) magnet. Each magnet particle has a diameter of 15 mm and a height of 20 mm. The arrangement of pairs between particles and the relative position of mice during exposure to MMF are shown in Fig. 1A and 1B. The magnetic induction intensity on the device and 1cm away from the device was measured by MIST magnetic field space-time imaging system (Eastforce Superconducting Technology Co., Ltd, Beijing, China), and ranged from 20 mT to 100 mT (Fig. 1C). The direction of magnetic induction lines is shown in Fig. 1D.

**Figure 1.**
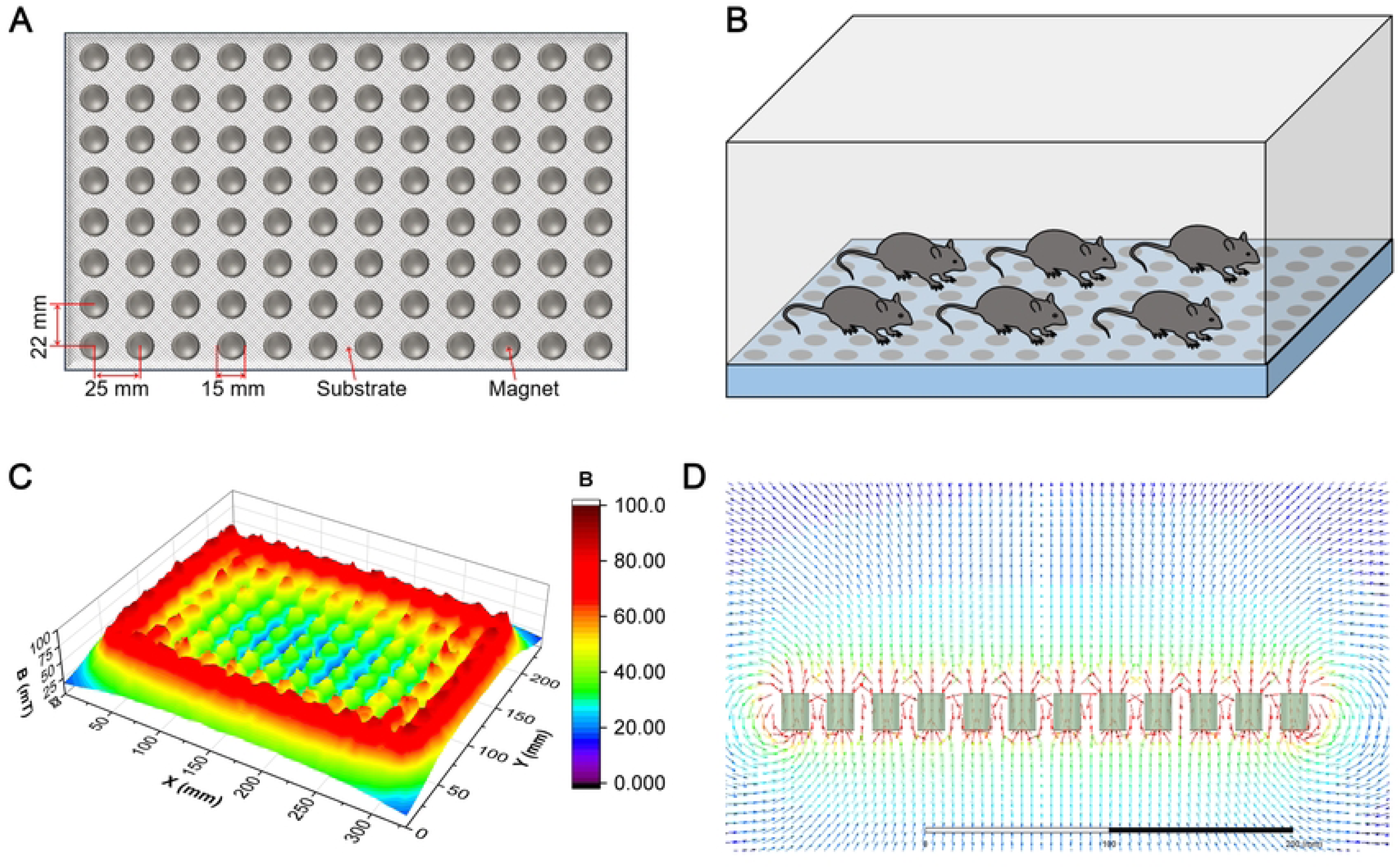
Moderate static magnetic field (MMF) exposure device for mice. (**A**) The diagram of magnetic plate that provide MMF. (**B**) Relative position of mice during exposure to MMF. (**C**) Magnetic field distribution in the mouse exposure area, 1 cm above the magnetic plates. (**D**) Direction and distribution of magnetic induction lines in magnetic exposure systems.

### BMD and BMC Examination

After eight weeks treatment, all mice were anesthetized and placed in a Dual-energy X-ray scanner (InAlyzer; MEDIKORS, Korea) to measure bone mineral density (BMD) and bone mineral content (BMC), including total body, femur, tibia and lumbar vertebrae.

### Bone Microarchitecture Examination

The left femur and lumbar vertebrae of mice were used for bone microarchitecture detection by a high-resolution micro-CT scanner (VivaCT80, SCANCO Medical AG, Bassersdorf, Switzerland). Scanning was performed at 70 kV, 114μA, 250ms without a filter. During the microstructure analysis of femur, area-of-interest (ROI) of 1-mm-long from the distal growth plate 0.5 mm to 1.5 mm were defined for analysis of trabecular bone, and another 1-mm-long ROI from the distal growth plate 5 mm to 6 mm were selected for analysis of cortical. Meanwhile, the trabecular bone in the L4 lumbar vertebrae were analyzed to evaluate the impact of the MMF exposure on bone loss in lumbar vertebrae. For the assessment of vertebral trabecular bone, the ROI was established within 5 mm from the edge of the cortical bone.

The parameters of the ROI were three-dimensionally reconstructed using Scanco analysis software (SCANCO Medical AG, Bassersdorf, Switzerland) to analyze trabecular structural parameters including bone volume fraction (BV/TV), trabecular number (Tb.N), trabecular thickness (Tb.Th), trabecular separation (Tb.Sp), BMD; and cortical bone structural parameters including cortical thickness (Ct.Th), total cross-sectional tissue area (Tt.Ar), cortical area fraction (Ct.Ar/Tt.Ar), cortical porosity (Ct.Po), and tissue mineral density (TMD).

### Bio-mechanical Examination

The left tibia was placed on two support points spaced 10 mm apart in a material mechanics machine, and then the upper loading device was perpendicular to the center of the tibia and moved downward at a rate of 1 mm/min to complete the three-point bending test. The inner and outer diameters of the long and short axes of the tibial fracture surface were measured by Digital Microscope KH-8700 (HIROX, Japan). Then, the computer-generated load-displacement curves and the stress-strain curve were analyzed to assess the mechanical properties of the tibia, including stiffness, ultimate load, ultimate displacement, bending energy absorption, ultimate stress, ultimate strain, elastic modulus, and toughness.

Take the L3 lumbar vertebra, remove the soft tissue on both sides, and use axial compression to test the mechanical properties of the vertebrae. The loading device was positioned perpendicular to the vertebral cross-section and moved downward at a rate of 1 mm/min. Then, the computer-generated load-displacement curves and the stress-strain curve were analyzed to assess the mechanical properties of the vertebrae, including stiffness, ultimate load, and elastic modulus.

### Histochemical Analysis

After the bone microstructure examination, the femur was collected and soaked in 10%EDTA with a PH of 7.5 and decalcified at 4 °C for two weeks, during which the decalcified fluid was changed every three days. The decalcified femur was embedded in paraffin and cut into 5mm thick sections using a semi-automatic rotary microtome (Leica Biosystems RM2245, Germany). Osteoblasts were observed by hematoxylin and eosin staining (H&E; Beyotime Biotechnology, Shanghai, China). Osteoclasts were identified by tartrate-resistant acid phosphatase staining (TRAP; Sigma-Aldrich, USA). The images of bone sections were acquired by Nikon 80i microscope (Nikon, Japan). Osteoblastogenesis was evaluated by analyzing osteoblast number per bone surface (N.Ob/BS), and osteoclastogenesis was estimated by quantizing osteoclast number per bone surface (N.Oc/BS) and osteoclast surface per bone surface (Oc.S/BS) by using Image-J software.

### Biochemical Analysis

The blood samples were collected through cardiac puncture and centrifuged at 1000g for 10 min to obtain serum. Bone turnover markers (BTMs), including propeptide of type I procollagen (PINP), osteocalcin (OCN), and beta-isomer of the C-terminal telopeptide of type I collagen (β-CTX) were detected by ELISA kit (JiangLai biological, Shanghai, China). All procedures are performed in accordance with the kit instructions.

### Clinical Trial Design

A randomized single-blind trial was conducted in People’s Hospital of Longhua between July 21^st^ 2021 and August 31^st^ 2023. This clinical trial was performed in accordance with the approved institutional ethical protocol (Ethical Review of People’s Hospital of Longhua [2021] No.106) and registered in the “Chinese Clinical Trial Registry (ChiCTR)” (ChiCTR2100048604). All procedures will be performed according to the Declaration of Helsinki. All individuals were signed the informed consent from providing their written consent prior to participation after being advised, verbally and written, of the objectives, risks, and benefits of the study.

#### Participants

Postmenopausal women with osteoporosis (diagnosed by dual energy X-ray absorptiometry, T score ≤ −2.5), aged between 55 and 70 years, were recruited in this study by advertisement, self-referral, or physician referral at People’s Hospital of Longhua (Shenzhen, China). Patients with the following criteria were excluded: (i) Prior spine surgery; (ii) Had taken fracture within 6 months before enrolment; (iii) Had taken Glucocorticoids, estrogen, or diuretic drugs within 6 months before enrolment; (iv) Had taken drugs affecting bone metabolism within 6 months before enrolment, excluding calcium agents and ordinary vitamin D; (v) Other diseases that affect bone metabolism, including thyroid disease, osteomalacia, rheumatoid arthritis, tumor, cushing’s disease, diabetes, etc.; (vi) Uterus or ovaries removed; (vii) Heart disease, or implantable pacemaker wearer; (viii) BMI ≥ 28; (ix) Hematological system diseases; (x) Abnormal liver and kidney function (Deviation of indicators by more than twice the normal value); (xi) Alcoholism or drug abuse.

Sample size was calculated (G*Power 3.1.9, Christian-Albrechts-Universitat, Kiel, Germany) using pain during areal bone mineral density (aBMD) as the primary outcome, α = 0.05, and power of 80% for between-group analysis. Considering a mean difference of 0.1 and a standard deviation of 0.07 in the mean aBMD between groups, sample size was estimated as 9 individuals per group. However, to account for losses during the follow-up period, the final sample size was 30 participants (15 per group).

#### Interventions

The participants were randomized into two groups: Control group and MMF group. Specifically, a series of random numbers were generated on SPSS software with a seed number set by a statistician (who did not participate in the inclusion of cases and experimental research processes), where every 2 random numbers formed a block. The participants were assigned to the corresponding groups in chronological order of enrollment. The grouping table is kept by the statistician and strictly confidential. Participants will be blinded since they will not know the exercise performed in the other group, and interventions will be performed individually. Participants in the MMF group were instructed to wear a waist belt containing the MMF device (Fig. 2) for 6 hours each day, five days a week. The wearable MMF device was composed of NdFeB, with the same array as the MMF exposure device for mice, which ensured that the participants’ lumbar vertebrae (L1-L4) exposed to MMF (>1 mT) during the wearing process (Fig. 2C-E). The relative position between the device and the participants during the wearing process was showed as Fig 2F. To avoid interference from placebo effects, the belt without MMF have been conducted, with the appearance, material, and weight same as wearable MMF device. Participants in the control group were instructed to wear the waist belt without MMF for 6 hours each day, five days a week. All participants received calcium (1200 mg), calcitriol (50 μg), and salmon calcitonin (50 μg) once daily as a basic treatment. The entire experimental process lasted for 90 days. The workflow of study was outlined in Fig. 3. Clinical evaluation was performed by orthopaedic surgeons.

**Figure 2.**
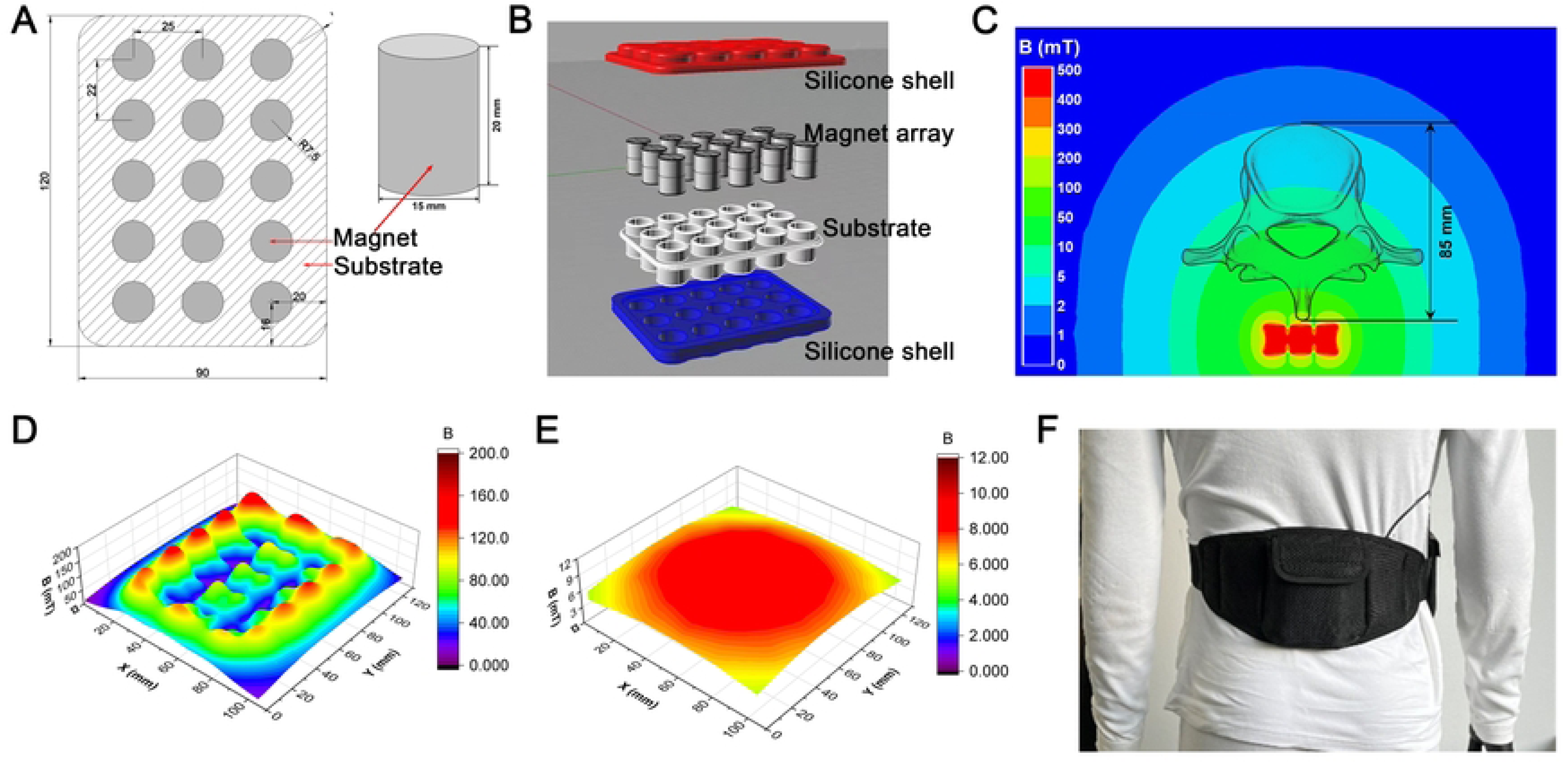
Moderate static magnetic field (MMF) device in clinical trial. (**A**) Schematic diagram of magnet size and array layout of MMF device. (**B**) Magnetic packaging structure. (**C**) The magnetic field distribution of vertebrae during MMF exposure was simulated using ANSYS software. (D, E) Magnetic field distribution at 5 mm (**D**) and 90 mm (**E**) above the magnet were measured by MIST magnetic field space-time imaging system. (**F**) Diagram of the body-worn MMF device.

**Figure 3.**
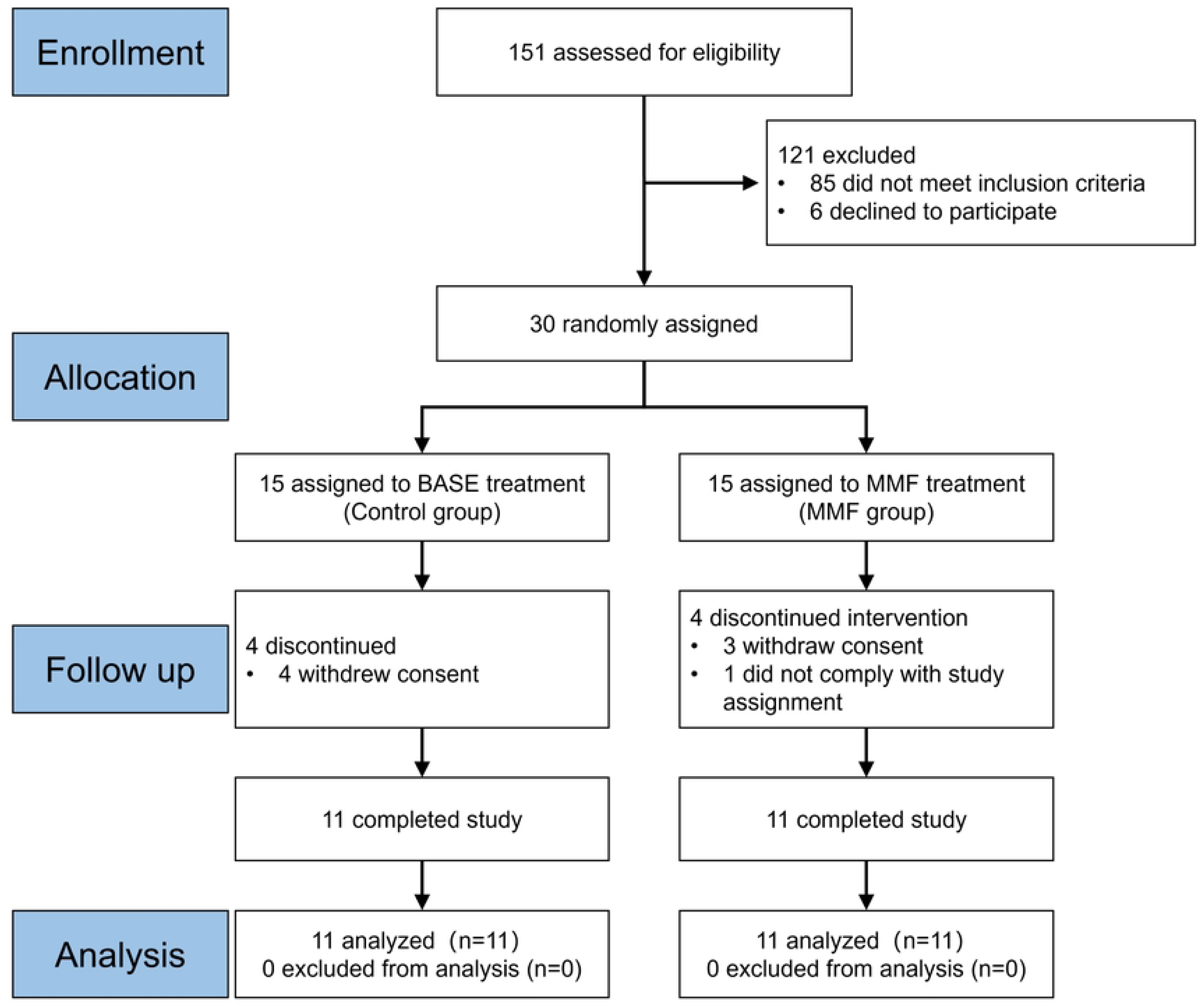
Flowchart of the clinical trial.

### Procedures

Participants’ characteristics (age, body mass, height, body mass index, and age of menopause) were collected at baseline assessment. Primary and secondary outcomes were measured at the baseline assessment, immediately after the 90 days intervention period. The primary outcomes was areal bone mineral density (aBMD). The secondary outcomes were serum bone turnover markers levels, and the Visual Analogical Scale (VAS) of Low Back Pain.

#### BMD Analysis

aBMD of the lumbar spine, total hip, and femoral neck were assessed at 0 and 3 months by dual x-ray absorptiometry (DXA) using a Hologic QDR 4500A densitometer (Hologic, Waltham, MA, USA). The coefficients of variation for DXA repeated measurements of the lumbar spine, total hip, and femoral neck were 0.7%, 1.4%, and 1.9%, respectively.

#### Biochemical Analysis

Fasting morning blood samples were obtained at 0 and 3 months. Serum bone formation markers P1NP and OCN as well as the bone resorption marker β-CTX were measured using the Au-400 automated biochemical analyzer (Olympus, Tokyo, Japan).

#### Visual Analogical Scale (VAS) of Low Back Pain

The participants were asked to express the intensity of the perceived low back pain (lumbar pain) on a one-dimensional scale. The scale was divided into 10 equal segments to represent distinct levels of pain. At one end, labeled as 0, participants indicated the absence of pain, while at the other end, labeled as 10, they expressed the highest imaginable level of pain[21].

### Monitoring System of Wearable State of MMF Device

To evaluate the daily wearing time of the participants during the clinical trial, a corresponding real-time monitoring and statistical system was constructed. The hardware components of the monitoring system consisted of sensors, signal generation module with power supply (Fig. S1A and B). The sensors were capable of collecting information about the wearing state at intervals of every 15 seconds, including parameters such as temperature and respiration. Subsequently, the sensors transmitted the collected information to the signal generation module, further uploading it to the cloud-based monitoring system’s backend software utilized 4G signals (Fig. S1D). Researchers were able to observe the participants’ wearing status of the device in real-time through the monitoring cloud platform and perform statistical analysis on wearing time (Fig. S1E and F). The wearable device monitoring system was not only designed to enhance data validity, but also to enable researchers to remind participants about wearing the devices, thereby promoting participant compliance.

### Statistical analysis

The normally distributed continuous data will be expressed as mean ± standard deviation (SD), as median (IQR) for endpoints without a normal distribution. The data about mean aBMD percentage change in clinical will be expressed trial as mean ± standard error (SE), as per the reporting conventions for this study area. The normal distribution was tested by Kolmogorov-Smirnov test with *P*>0.10. The comparison of group differences in normally distributed continuous data was performed using t-tests or ANOVA. Non-parametric rank-based tests were employed for the comparison of group differences in non-normally distributed continuous data. The comparison of group differences in categorical data was conducted using the chi-square test, and Spearman’s rank correlation analysis was employed to assess the correlation between variables. *P*<0.05 will be considered to be statistically significant. All the statistical data will be analyzed by GraphPad Prism (version 8; GraphPad Software, Inc., San Diego, CA, USA).

## Results

### Effects of MMF exposure on bone quality in OVX mice

The body weight of mice was measured every four days, and there was no statistically significant difference in body weight among the different groups (Fig. S2). The effects of MMF exposure on BMD and BMC in OVX mice were detected by DXA and shown in table S1. The results showed that the OVX decreased the BMD and BMC in mice. The MMF exposure increased BMD of the total body, femur, and lumbar vertebrae in OVX mice. Meanwhile, the femoral BMC and lumbar vertebral BMC in OVX mice were significantly increased by MMF exposure. While, there was no significant difference in the tibial BMD, total BMC, and tibia BMC in OVX-MMF group compared to OVX group.

Micro-CT scanning of the femur was used to evaluate effects of MMF exposure on the microstructure of weight-bearing bones in OVX mice. Representative three-dimensional imaging of cortical bone and trabecular bone were illustrated (Fig. 4A and B). In cortical bone of femoral mid-shaft, the results showed OVX significant decreased Ct.Th, Ct.Ar, Ct.Ar/Tt.Ar and TMD, while markedly increased Ct.Po. Furthermore, Ct.Ar/Tt.Ar was significantly increased in OVX-MMF group compared to the OVX group (Fig. 4C). For the trabecular bone of distal femur, OVX group showed significant decreases in BV/TV, Tb.N, Tb.Th and BMD, whereas significant increases in Tb.Sp. compared with the control group. Compared to the OVX group, MMF exposure significantly increased BV/TV, Tb.N, Tb.Th, and BMD, while decreased Tb.Sp in trabecular bone of the distal femur (Fig. 4D). In conclusion, these data suggested that MMF exposure ameliorates OVX-induced structural destruction of cortical bone and trabecular bone in the femur. Micro-CT scanning on the lumbar vertebra was used to evaluate the effect of MMF exposure on the microstructure of non-weight-bearing bones. Representative three-dimensional imaging of lumbar vertebra was illustrated in Fig. S3A. The statistical results showed that OVX decreased the BMD, BV/TV, Tb.N, and Tb.Th, while increased the Tb.Sp in mice. MMF exposure increased the BV/TV, Tb.Th, and BMD, while decreased the Tb.Sp of lumbar vertebra in OVX mice (Fig. S3B).

**Figure 4.**
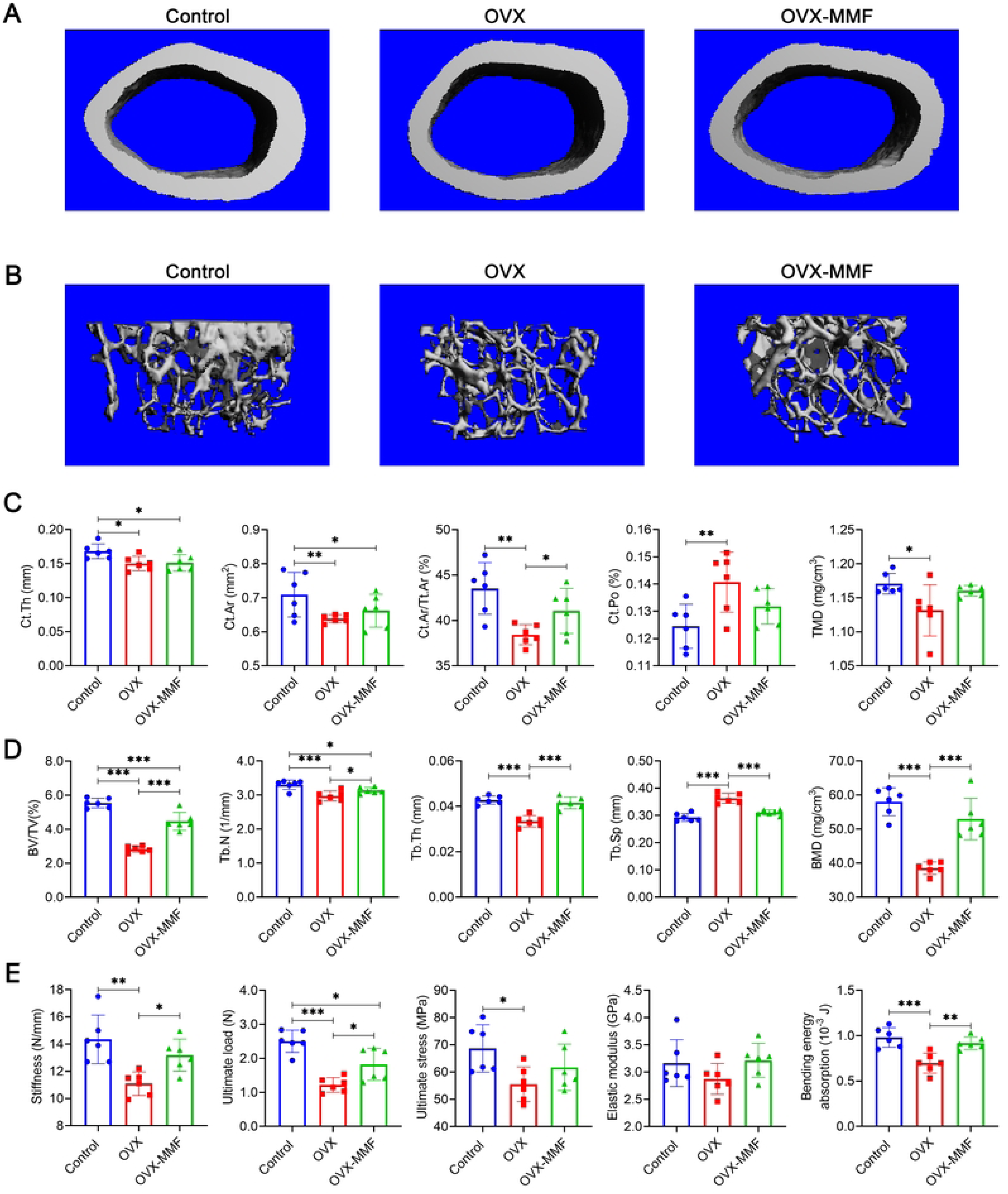
Effects of MMF exposure on bone structure and mechanical properties in OVX mice. **(A)** Three-dimensional images of cortical architecture in femoral mid-shaft by micro-CT scanning. **(B)** Three-dimensional images of trabecular architecture in distal femur by micro-CT scanning. **(C)** Structural parameters of cortical bone, including Ct.Th, Ct.Ar, Ct.Ar/Tt.Ar, Ct.Po, and TMD. **(D)** Structural parameters of trabecular bone, including BV/TV, Tb.N, Tb.Th, Tb.Sp, and BMD. **(E)** Mechanical properties of the tibia in mice were detected through the three-point bending test, including stiffness, ultimate load, ultimate stress, elastic modulus, and bending energy absorption. n =6. \**P*<0.05, \*\**P*<0.01, \*\*\**P*<0.001.

Three-point bending was used to evaluate changes in the mechanical properties of the tibia. The results exhibited stiffness, ultimate load, ultimate stress, elastic modulus, and bending energy absorption were significantly decreased in OVX group. Compared with the OVX group, the MMF exposure restored the stiffness, ultimate load, and bending energy absorption of tibia. Whereas, there was no significant differences in ultimate stress and elastic modulus between mice in OVX group and OVX-MMF group (Fig. 4E). The axial compression test was used to evaluate changes in the mechanical properties of the lumbar vertebra. The results showed that stiffness, elastic modulus, and ultimate load of lumbar vertebra were significantly reduced after OVX, but MMF exposure significantly increases the values of these parameters (Fig. S3C). These results suggested that MMF exposure significantly repairs the weakening of bone strength caused by OVX.

### Effects of MMF exposure on bone turnover in OVX mice

Bone formation is mainly mediated by osteoblasts, and bone resorption is mainly mediated by osteoclasts. The osteoblastogenesis and osteoclastogenesis were detected by H&E and TRAP staining in femur. The distribution of osteoblasts on the surface of cortical bone and trabecular bone were displayed in Fig. 5A. The statistical results showed OVX decreased the N.Ob/BS of trabecular bone and cortical bone. Statistical comparisons demonstrated that MMF exposure for 8 weeks significantly increased the N.Ob/BS of trabecular bone, while have no significant effect on the N.Ob/BS of cortical bone compared with the OVX mice without MMF exposure (Fig. 5B, C). The OVX treatment reduced the levels of OCN and P1NP in the serum of mice, and the MMF exposure increased the P1NP content in OVX mice (Fig. 5D, E). The distribution of osteoclasts on the surface of cortical and trabecular bone were displayed in Fig 5F. Statistical results demonstrated OVX increased the N.Oc/BS and OC.S/BS of trabecular bone and cortical bone. MMF exposure could significantly decreased the N.Oc/BS and OC.S/BS of cortical and trabecular bones in OVX mice (Fig. 5G-J). Serum biochemical analysis showed that MMF exposure significantly decreased bone resorption markers β-CTX compared with the OVX group (Fig. 5K).

**Figure 5.**
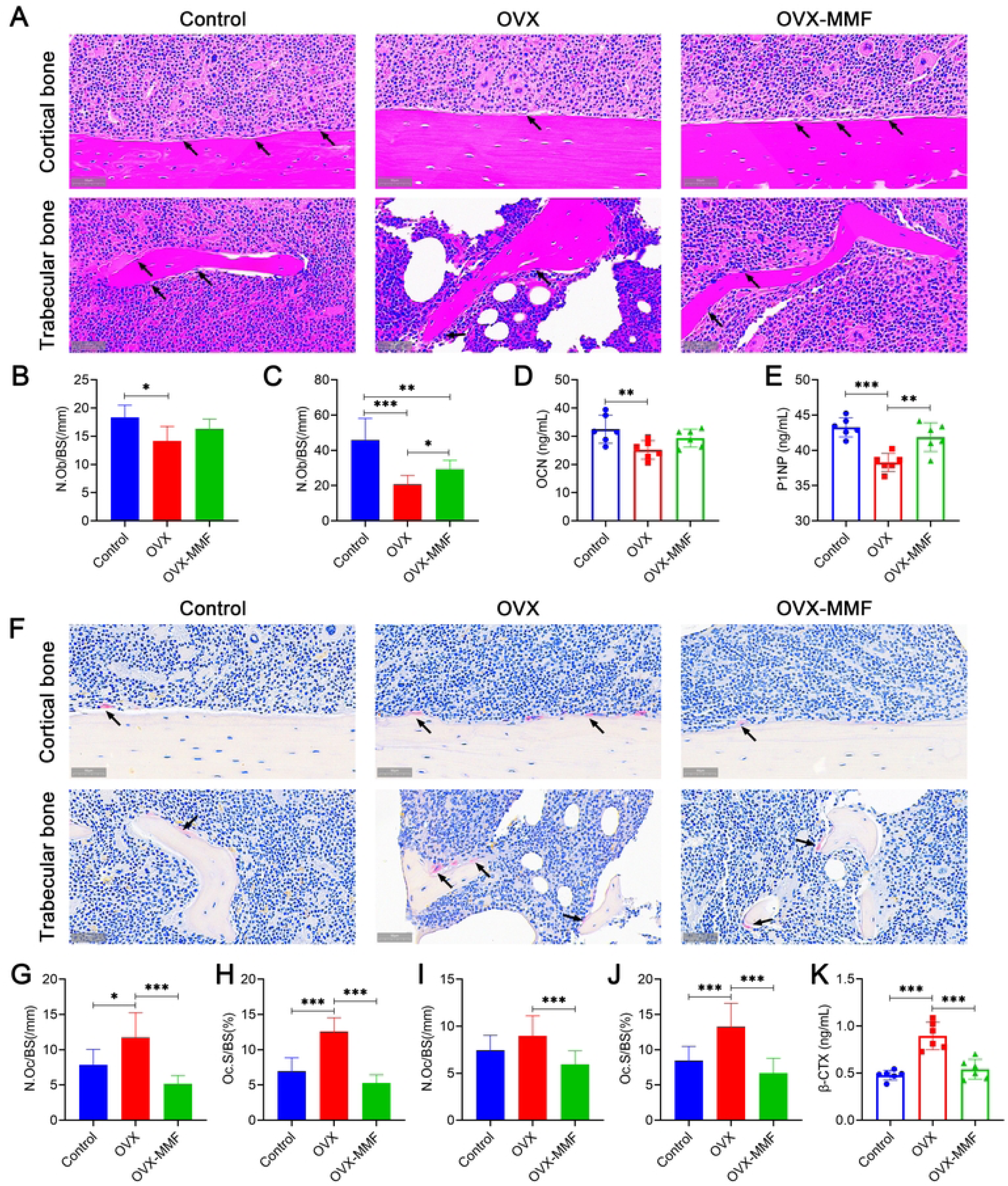
Effects of MMF exposure on bone remodeling in OVX mice. **(A)** H&E staining of cortical bone in femoral mid-shaft and trabecular bone in distal femur (Scale bar = 100 μm). Black arrows mark osteoblasts. **(B)** The osteoblast number per bone surface (N.Ob/BS) in cortical bone. **(C)** The osteoblast number per bone surface (N.Ob/BS) in trabecular bone. Serum bone formation markers, OCN **(D)** and P1NP **(E)** were detected using the commercial ELISA kits. **(F)** TRAP staining of cortical bone in femoral mid-shaft and trabecular bone on distal femur (Scale bar = 100 μm). Osteoclast number per bone surface (N.Oc/BS) **(G)** and osteoclast surface per bone surface (Oc.S/BS) **(H)** in cortical bone. osteoclast number per bone surface (N.Oc/BS) (**I**) and osteoclast surface per bone surface (Oc.S/BS) (**J**) in trabecular bone. (**K**)Serum bone resorption marker, β-CTX was detected using the commercial ELISA kit. n =6. \**P*<0.05, \*\**P*<0.01, \*\*\**P*<0.001.

### Baseline characteristics of subjects in clinical trial

A RCT was implemented to initially and exploratively evaluate the efficacy of MMF in postmenopausal osteoporosis. As shown in Figure 3, 30 subjects were ultimately enrolled in the clinical trial. During the course of the trial, 8 cases withdrew from the trial, of which 7 cases (4 cases in the control group and 3 cases in the MMF group) voluntarily withdrew their consent and 1 case (in the MMF group) did not comply with the study procedures. No participants reported any adverse events throughout the clinical trial. The baseline characteristics of the 22 subjects who persisted to the trial endpoint were showed in Table 1. Participants in both groups had similar baseline characteristics, including age, age at menopause, height, weight, BMI, serum BTMs, and VAS scores of low back pain.

**Table 1.**
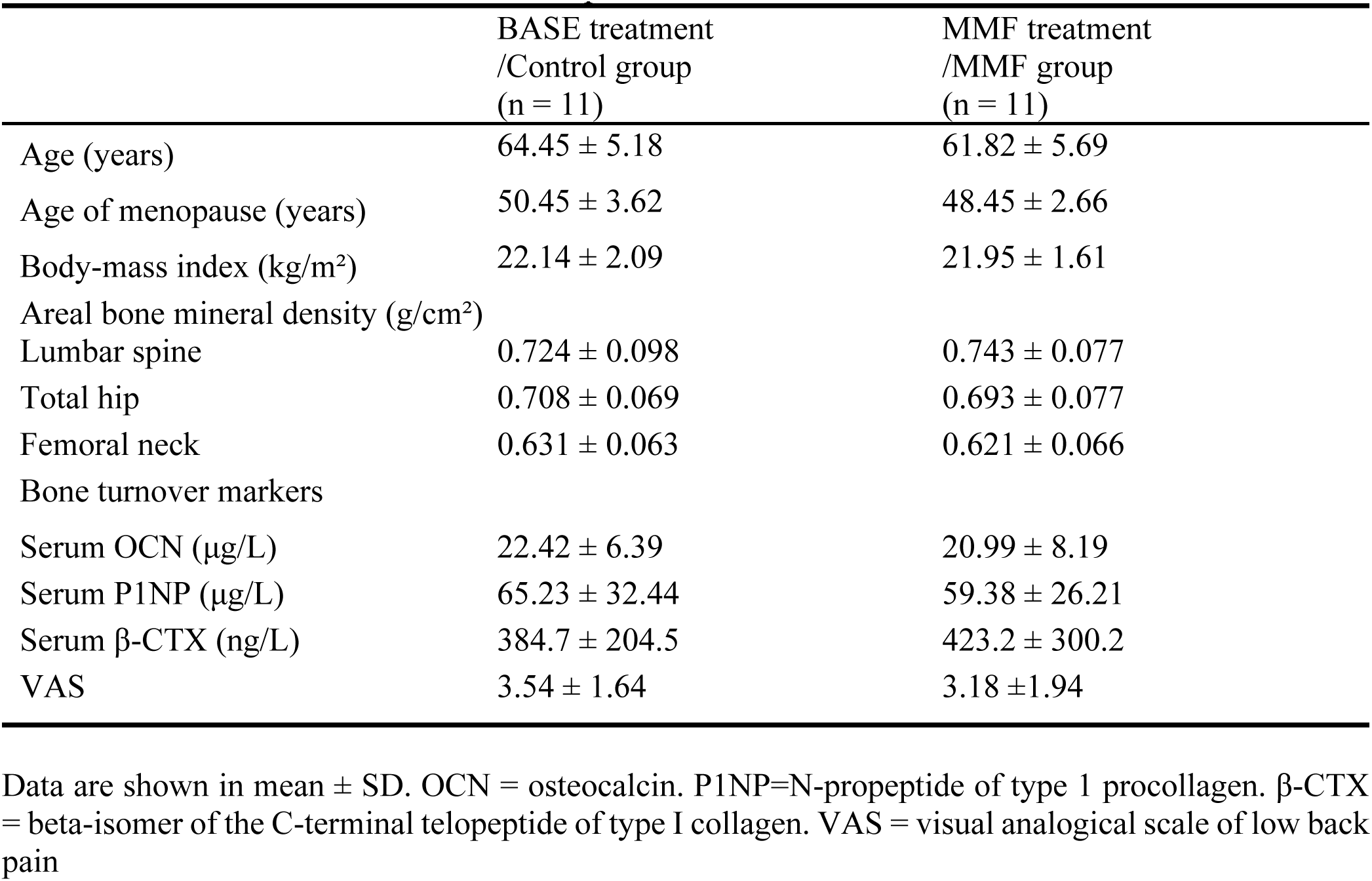
Participants Baseline Characteristics.

|  | BASE treatment<br>/Control group<br>(n = 11) | MMF treatment<br>/MMF group<br>(n = 11) |
| --- | --- | --- |
| Age (years) | 64.45 ± 5.18 | 61.82 ± 5.69 |
| Age of menopause (years) | 50.45 ± 3.62 | 48.45 ± 2.66 |
| Body-mass index (kg/m<sup>2</sup>) | 22.14 ± 2.09 | 21.95 ± 1.61 |
| Areal bone mineral density (g/cm<sup>2</sup>) |  |  |
| Lumbar spine | 0.724 ± 0.098 | 0.743 ± 0.077 |
| Total hip | 0.708 ± 0.069 | 0.693 ± 0.077 |
| Femoral neck | 0.631 ± 0.063 | 0.621 ± 0.066 |
| Bone turnover markers |  |  |
| Serum OCN (µg/L) | 22.42 ± 6.39 | 20.99 ± 8.19 |
| Serum P1NP (µg/L) | 65.23 ± 32.44 | 59.38 ± 26.21 |
| Serum β-CTX (ng/L) | 384.7 ± 204.5 | 423.2 ± 300.2 |
| VAS | 3.54 ± 1.64 | 3.18 ± 1.94 |

Data are shown in mean ± SD. OCN = osteocalcin. P1NP=N-propeptide of type 1 procollagen. β-CTX = beta-isomer of the C-terminal telopeptide of type I collagen. VAS = visual analogical scale of low back pain

### Effects of MMF wearable device treatment on bone mineral density of subjects in clinical trial

After 90 days treatment, the aBMD of lumbar spine, total hip, and femoral neck of participants were tested by DXA and showed in table S2. The statistical data showed that the lumbar spine aBMD had increased by 1.88% (SE 1.39) from baseline in the control group, by 2.86% (SE 1.41) in the MMF group (Fig. 6A). The total hip aBMD had increased by 0.09% (SE 1.11) from baseline in the control group, by 2.37% (SE 1.09) in the MMF group (Fig. 6B). The femoral neck aBMD had increased by 1.29% (SE 1.69) from baseline in the control group, by 2.96% (SE 1.27) in the MMF group (Fig. 6C). However, there were no statistically significant changes on aBMD at the lumbar spine, total hip, and femoral neck of participants in MMF treatment (MMF group) compared with non-MMF exposure (control group). Furthermore, we analyzed the relationship between the lumbar aBMD of participants and their age. The enhancing effect of MMF treatment on aBMD had a higher negative correlation with participants’ age compared to the control group (Fig S4).

**Figure 6.**
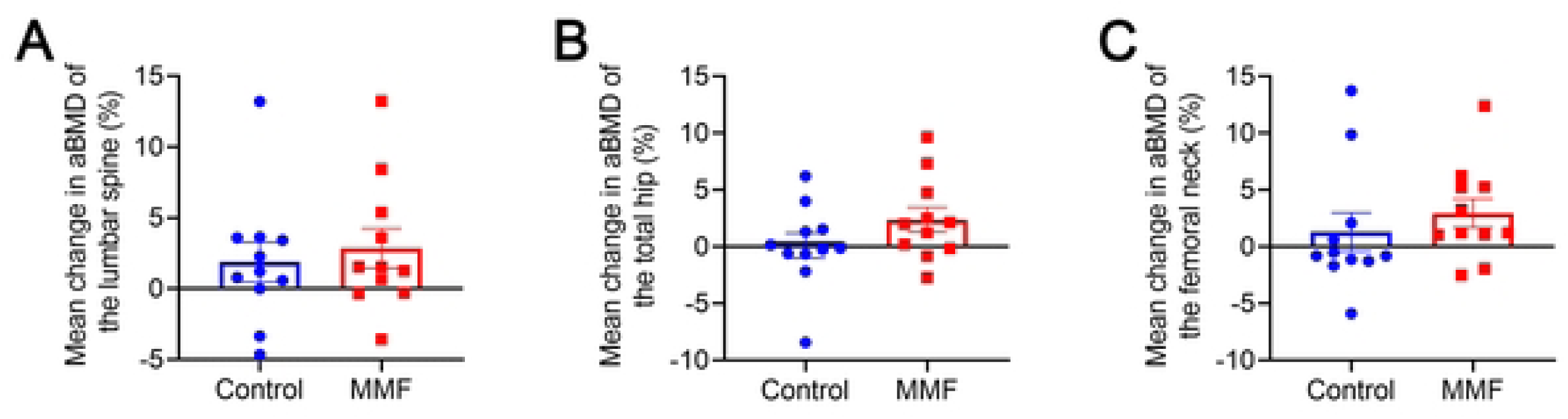
Mean percentage change in aBMD of the lumbar spine (A), total hip (B), and femoral neck (C) after 90 days treatment in clinical trial. aBMD=areal bone mineral density.

### Effects of MMF treatment on bone turnover markers and low back pain of subjects in clinical trial

After 90 days treatment, BTMs level of participants were showed in table S2. Compared to the control group, levels of the bone formation marker OCN decreased more in the MMF group (−22.87% *vs* −6.58%, *P* = 0.0135) (Fig. 7A). The levels of P1NP showed a lesser decrease in MMF group than the control group (−14.54% *vs* −31.86%, *P* = 0.0096) (Fig. 7B). The level of bone resorption marker β-CTX increased by 4.85% from the baseline value in the control group (Fig. 7C). In contrast, β-CTX levels decreased by 38.83% from the baseline value in the MMF group. Furthermore, the rate of β-CTX change in the MMF group showed a significant difference when compared to the control group (*P* = 0.0377).

**Figure 7.**
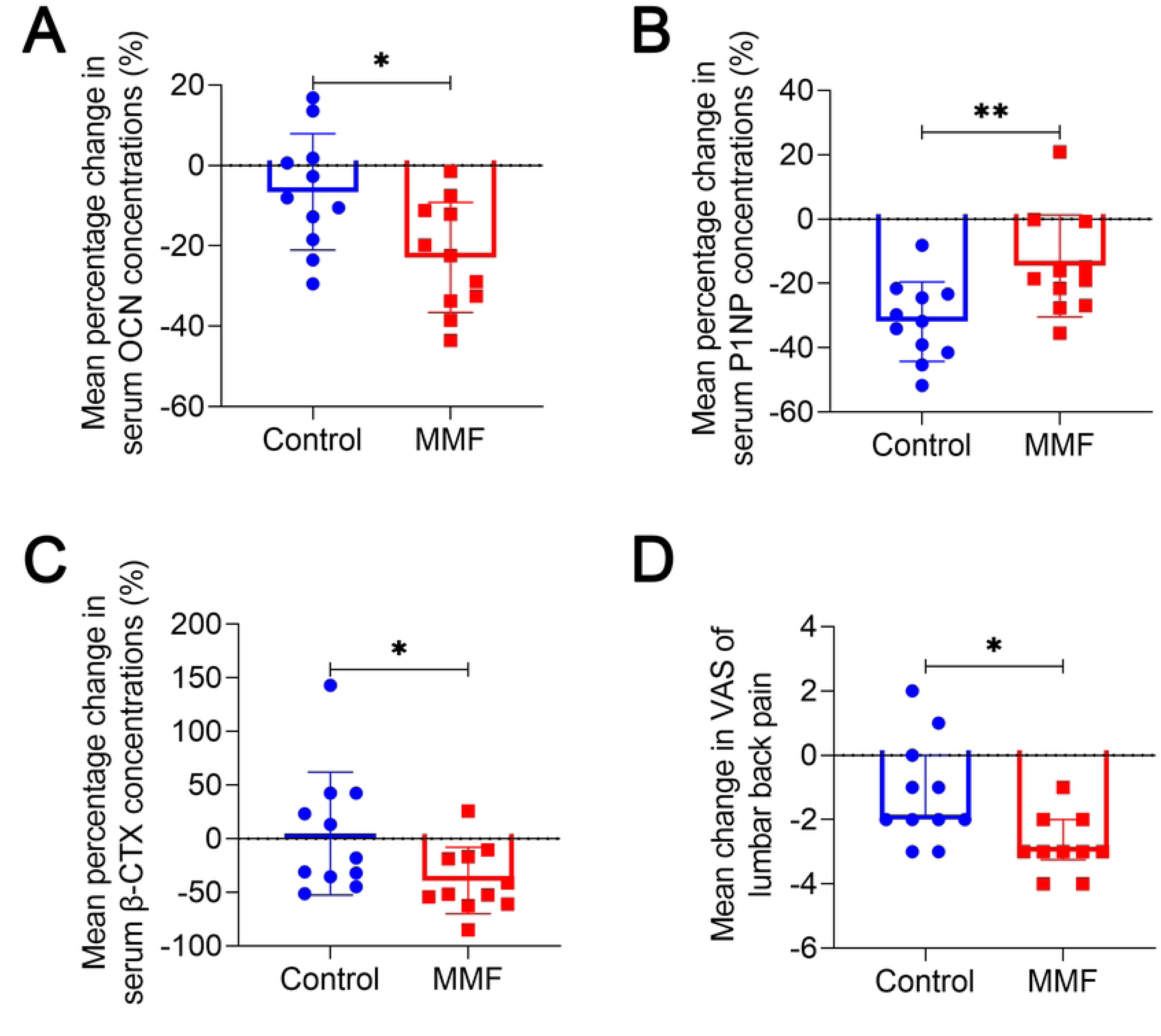
Change in bone turnover markers and VAS of low back pain after 90 days treatment in clinical trial. (A-C) Mean percentage change in serum concentrations of bone formation makers, OCN (A) and P1NP (B), as well as bone resorption marker β-CTX (C). (D) Mean change in VAS of low back pain after 90 days treatment in clinical trial. OCN= osteocalcin. P1NP=N-terminal propeptide of type 1 procollagen. β-CTX= beta-isomer of C-Terminal Telopeptide of Type I Collagen. \**P*<0.05, \*\**P*<0.01.

Due to the presence of symptoms of low back pain among some participants, the VAS score was employed to assess the participants’ low back pain before and after the treatments. The results showed that VAS scores were significantly lower in the MMF group compared to the control group (−2.800 *vs* −1.182, *P* = 0.0197), suggesting that MMF treatment relieved lower back pain of participants (Fig. 7D).

## Discussion

MMFs have been used in orthopedics diseases studies for over half a century. In this study, we performed a first clinical trial to evaluate the effects of MMF on PMOP. Although this study was a small sample size RCT, the preliminary results demonstrated the effectiveness of MMF exposure for PMOP therapy, including reduced bone turnover and alleviated lower back pain.

As a chronic and progressive disease, the treatment of PMOP typically requires several years and may even extend throughout a patient’s lifetime[22, 23]. Therefore, the compliance of patients is crucial for the effectiveness of osteoporosis therapy[24, 25]. In order to improve patient compliance, we conducted a wearable MMF device which could be carried by the patients themselves, and allowing them to complete the trial in their own homes. However, home-based trial without the presence of researchers cannot guarantee participants’ adherence to the designated trial procedures. Therefore, sensors were incorporated into the device to monitor the participants’ wearing status in real-time.

In this study, the continuous MMF exposure for 8 weeks significantly increased BMD and microstructure of the femur and lumbar vertebrae in OVX mice. The participants in control and MMF group demonstrated a well compliance, and no adverse reactions were observed in clinical trial. Meanwhile, the results indicated that although there was no statistically significant difference in the improvement of BMD of lumbar spine between the MMF group and the control group, the MMF treatment demonstrated a greater magnitude of increase in BMD in PMOP patients. Clearly, expand the sample size of clinical trial may lead to similarly significant effects, as observed in the effects of MMF exposure on bone mass in OVX mice. Furthermore, this discrepancy may be attributed to the relatively short duration of the clinical trial. Therefore, the future clinical research on the effects of SMFs on PMOP should consider designing longer trial periods to further investigate the potential impact.

The results of clinical trial showed that all two treatments approaches exhibited a decrease in the concentration of bone formation and bone resorption markers in serum. The MMF treatment decreased the levels of serum β-CTX in both postmenopausal women and OVX mice. In comparison to the control group, the MMF wearable device reduced the extent of P1NP decrease in clinical trial. The improvement in bone turnover induced by the decreased of β-CTX levels and the increased of P1NP may contribute to the beneficial effects of MMF wearable device not only on the lumbar spine’s aBMD (exposed region) but also on the aBMD of total hip and femoral neck (non-exposed region) in participants. However, for another bone formation marker, OCN, MMF exposure did not significantly increase its serum levels in mice, even reduced serum OCN levels compared to the control group in clinical trials. This may be attributed to the inhibitory effect of MMF on osteoclast activity, as the circulating levels of biologically active OCN are regulated by osteoclasts. During the bone resorption process, the acidic environment created by osteoclasts leads to the conversion of inactive c-carboxylated osteocalcin (GlaOCN) to active undercarboxylated osteocalcin (GluOCN)[26]. The increased of osteoclast activity in mice resulted in elevated levels of GluOCN in the serum, and impaired osteoclast function could decreased the GluOCN level[27]. Regarding the fact that MMF exposure did not change serum OCN levels in OVX mice but decreased them in PMOP patients compared to their respective controls, which may be attributed to differences in MMF exposure patterns between animal and clinical study. It is worth noting that the clinical trial involved localized exposure to a MMF in the lumbar spine region, whereas in the animal experiment, mice were whole-body exposed to MMF. Furthermore, there are metabolic differences between mice and humans.

In this study, we also have investigated the correlation between age and BMD changes among participants in different groups. The results showed that there was minimal correlation between age and lumbar spine BMD changes in the control group. However, there was a negative correlation between lumbar spine BMD changes and age in MMF group, although no statistically significant. This suggests that MMF is more effective in improving BMD in younger postmenopausal osteoporosis (PMOP) patients. With the bone undergoes aging, the sensitivity of bone to mechanical stimuli was reduced gradually [28]. All subjects in SMFs are subject to the action of magnetic force, which is one of the primary mechanisms SMF biological effects[29, 30]. Therefore, the relationship between the effects of MMF on PMOP patients with different ages and the magnetic force deserves further research. Additionally, in the early postmenopausal period, the reduction in estrogen levels leads to rapid bone loss, with an annual bone loss rate of 3-5%. This accelerated bone loss can persist for 5-10 years and primarily affects trabecular bone. In the late postmenopausal period, the rate of bone loss slows down, and cortical bone loss becomes more predominant, and associated with an increased risk of fragility fractures[4]. Considering the significant regulatory effect of MMF on bone remodeling in the present study, it is possible that MMF may have a more pronounced therapeutic effect on bone in a period of rapid bone loss with high bone turnover. In the future, it would be interesting to include participants from different age groups to study the effects of MMF exposure on BMD at different stages of aging.

PMOP patients usually suffer from low back pain[31–33]. Various studies indicated that SMFs had an analgesic effects[34]. Suzan et al. found that 20 mT MMF exposure had a positive effect on patients with chronic lumbar radicular pain[35]. Studies indicated that the relief of chronic lumbar radicular pain by the SMF was associated with the magnetic field intensity[36, 37]. In this study, we found that 6 hour per day MMF treatment effectively alleviated low back pain in PMOP patients. This may be attributed to the fact that the relief of pain by the SMF is not only associated with the intensity of the magnetic field but also with the duration of the intervention. Due to the inconsistency of conclusions derived from a multitude of SMF treatments with varying intensities and exposure durations, further research is warranted to explore the relationship between the dosage of SMF exposure and its impact on osteoporosis and related pain.

However, this study still has several limitations. Firstly, the lack of in-depth mechanistic research. This is because the aim of this study primarily focused on exploring the clinical efficacy of MMF in treating PMOP. Our previous studies have demonstrated that SMFs regulated cellular and systemic iron metabolism, which is closely associated with osteoporosis[38–41]. Therefore, we will investigate the mechanisms of MMF therapy for PMOP based on iron metabolism in subsequent studies. Secondly, sample size of the clinical trial was relatively small. Since there were no clinical reference data on MMF for the treatment of osteoporosis, the initial intention of designing this clinical trial was to explore the therapeutic effects of MMF on PMOP using a small sample size as a preliminary investigation. Surprisingly, we found that MMF effectively regulated bone turnover, alleviated low back pain, and exhibited a tendency to increase lumbar spine BMD. Hence, we were eager to publish these preliminary results to promote the clinical application of MMF in the prevention and treatment of PMOP. Thirdly, the clinical trial in this study only had a single endpoint. The long-term therapeutic effects of MMF on osteoporosis or the potential long-term effects after MMF treatment cessation remain unclear. The multicenter, large-scale RCTs with multiple time points is necessary to comprehensively evaluate the therapeutic effects of MMF on osteoporosis. Furthermore, in this study, we have only investigated the effects of lumbar spine local MMF exposure on BMD and BTMs in PMOP patients. Therefore, further investigation is needed to explore the effects of whole-body MMF exposure or localized MMF exposure at other sites, such as the hip, on osteoporosis.

In conclusion, the MMF exposure not only improved the bone microstructure and bone remodeling homeostasis in OVX mice but also significantly enhanced the bone mechanical properties. The results from RCT demonstrated that 90-day MMF treatment significantly improved serum BTMs and relieved low back pain in PMOP patients. Furthermore, MMF treatment exhibited a positive effect on BMD of the lumbar spine in PMOP patients. It indicated that MMF is a potential adjuvant therapy for osteoporosis and will promote the clinical translation research and application of SMFs.

## Data Availability

All relevant data are within the manuscript and its Supporting Information files.

## Acknowledgements

This work was supported by the National Natural Science Foundation of China (52037007), the Shenzhen Science and Technology program (JCYJ20230807144159004), Chongming Project of the Heye Health Technology Co., Ltd (HY-I-202101), Space Medical Experiment Project of China Manned Space Program (HYZHXM01008), and Space Utilization Project of China Manned Space Program (YYWT-0901-EXP-04).

## Author contributions

SHW: Methodology, Data curation, Formal Analysis, Writing – original draft, Funding acquisition; JCY: Methodology, Data curation, Formal Analysis, Supervision, Writing –review & editing; YPW: Methodology, Software; CC: Data curation; SC: Data curation; YDW: Data curation; XL: Data curation; LLS: Data curation; XLL: Data curation; MG: Data curation, Formal Analysis; JHZ: Data curation; YWH: Data curation; WZ: Data curation; ZFG: Data curation; JCL: Data curation, Project administration; CLL: Data curation; XSB: Methodology; XLL: Methodology; LMD: Methodology, Software; THY: Methodology; ZQY: Methodology; MW: Methodology, Validation; YWF: Methodology; HZ: Conceptualization, Methodology, Project administration, Supervision, Validation; PS: Conceptualization, Methodology, Project administration, Supervision, Writing –review & editing, Funding acquisition.

---

**Figure S1.**
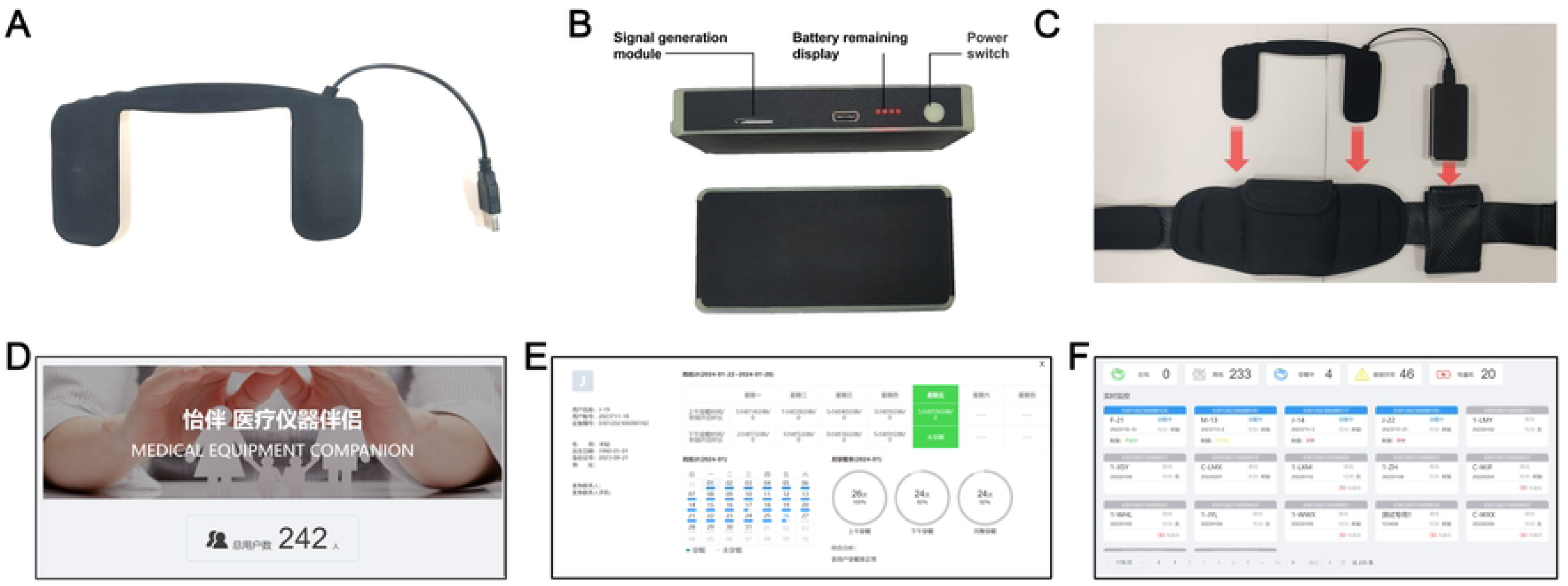
Wearable State Monitoring System of MMF device for clinical trial. (**A**) Sensor. (**B**) Signal generation module with power supply. (**C**) Schematic diagram of the location of the monitoring system hardware components on MMF device. (**D**) Homepage of cloud-based monitoring system’s backend software. (**E**) Real-time interface for displaying the wearing status of device. (**F**) Statistics screen for historical wear information.

**Figure S2.**
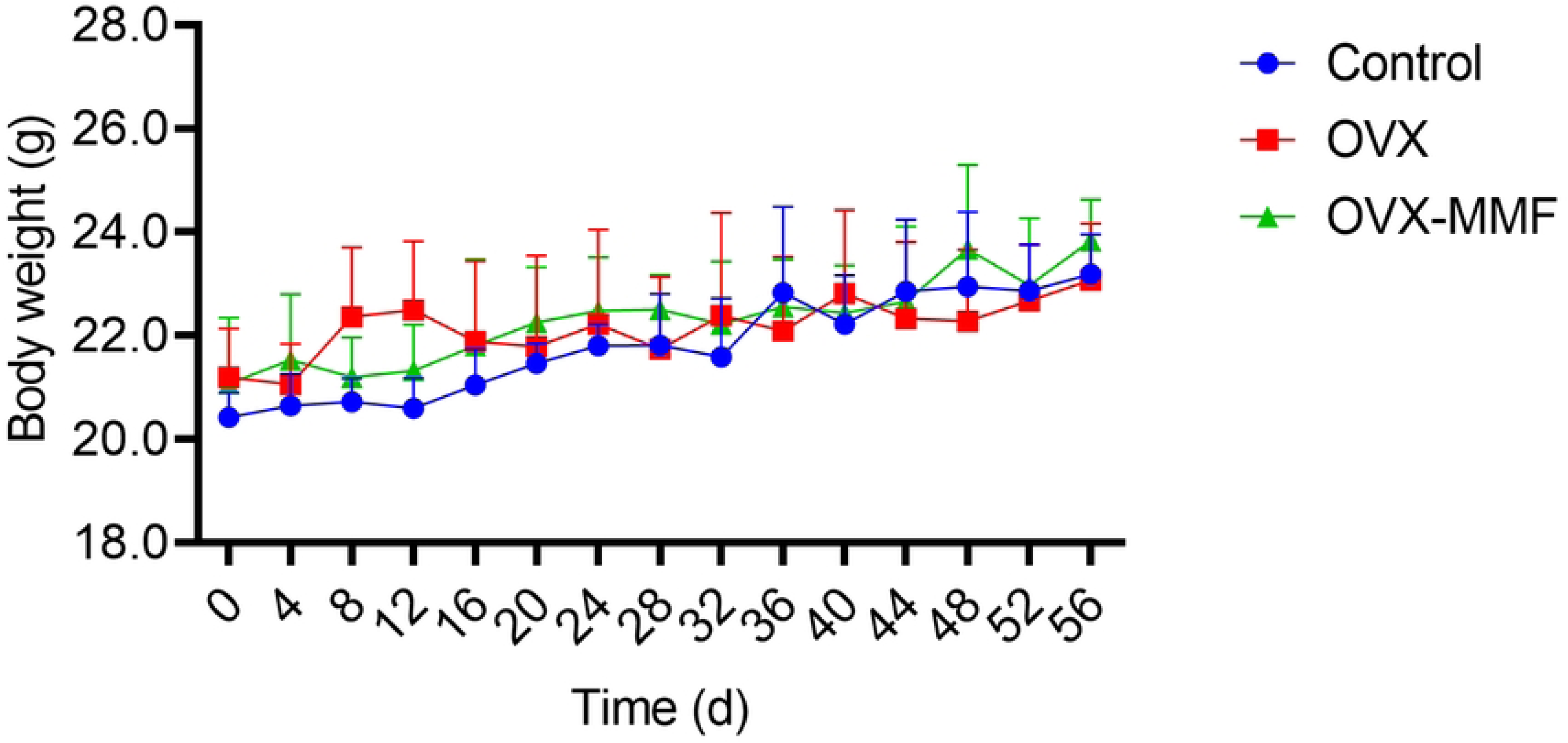
Effects of MMF exposure on body weight of mice.

**Figure S3.**
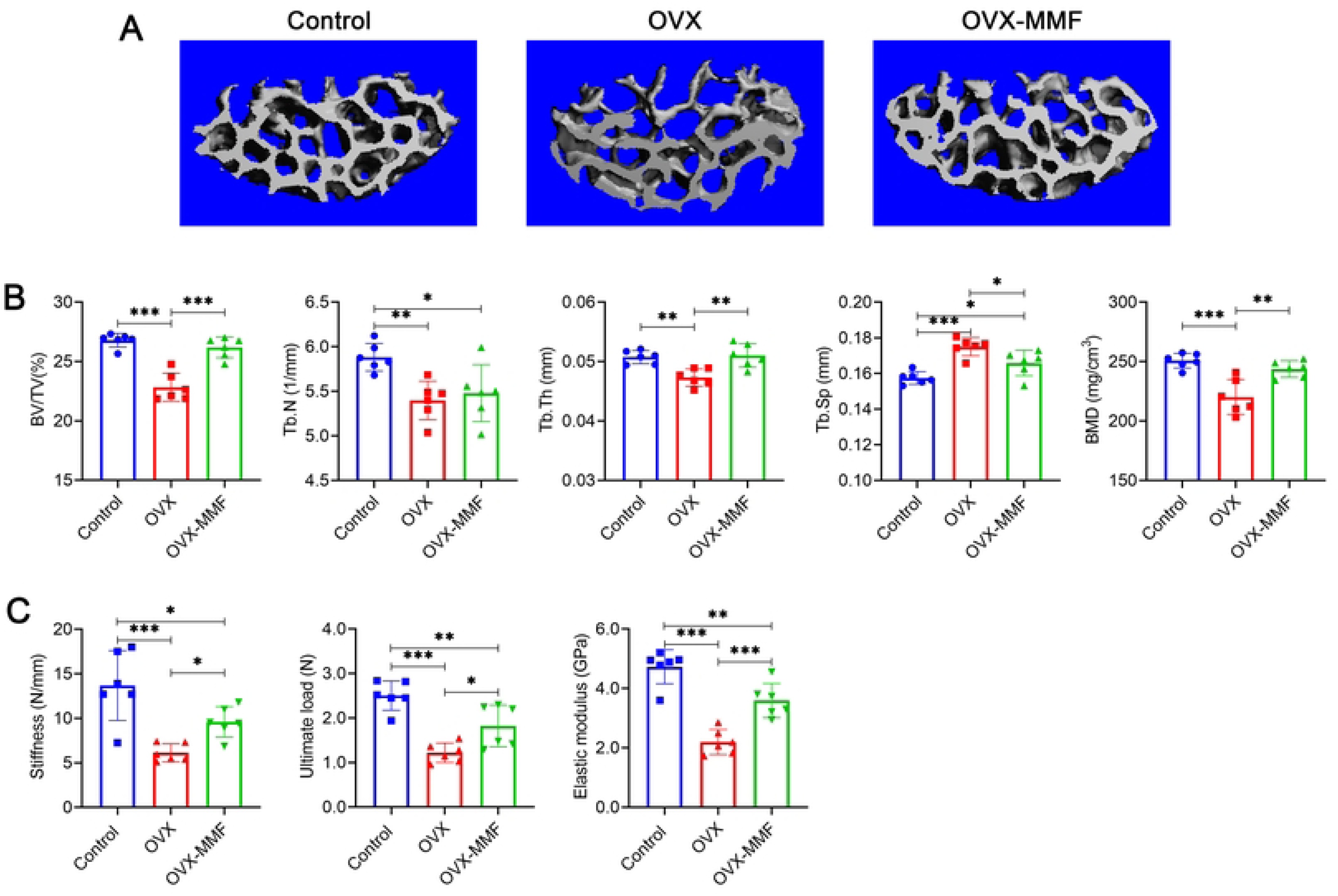
Effects of MMF exposure on microstructure and mechanical properties of the lumbar vertebrae in OVX mice. **(A)** Three-dimensional images of L4 lumbar vertebrae by micro-CT scanning. **(B)** Structural parameters of L4 trabecular bone, including BV/TV, Tb.N, Tb.Th, Tb.Sp, and BMD. **(C)** Mechanical properties of the L3 lumbar vertebrae in mice were detected through axial compression test, including stiffness, ultimate load, and elastic modulus. n =6. \**P*<0.05, \*\**P*<0.01, \*\*\**P*<0.001.

**Figure S4.**
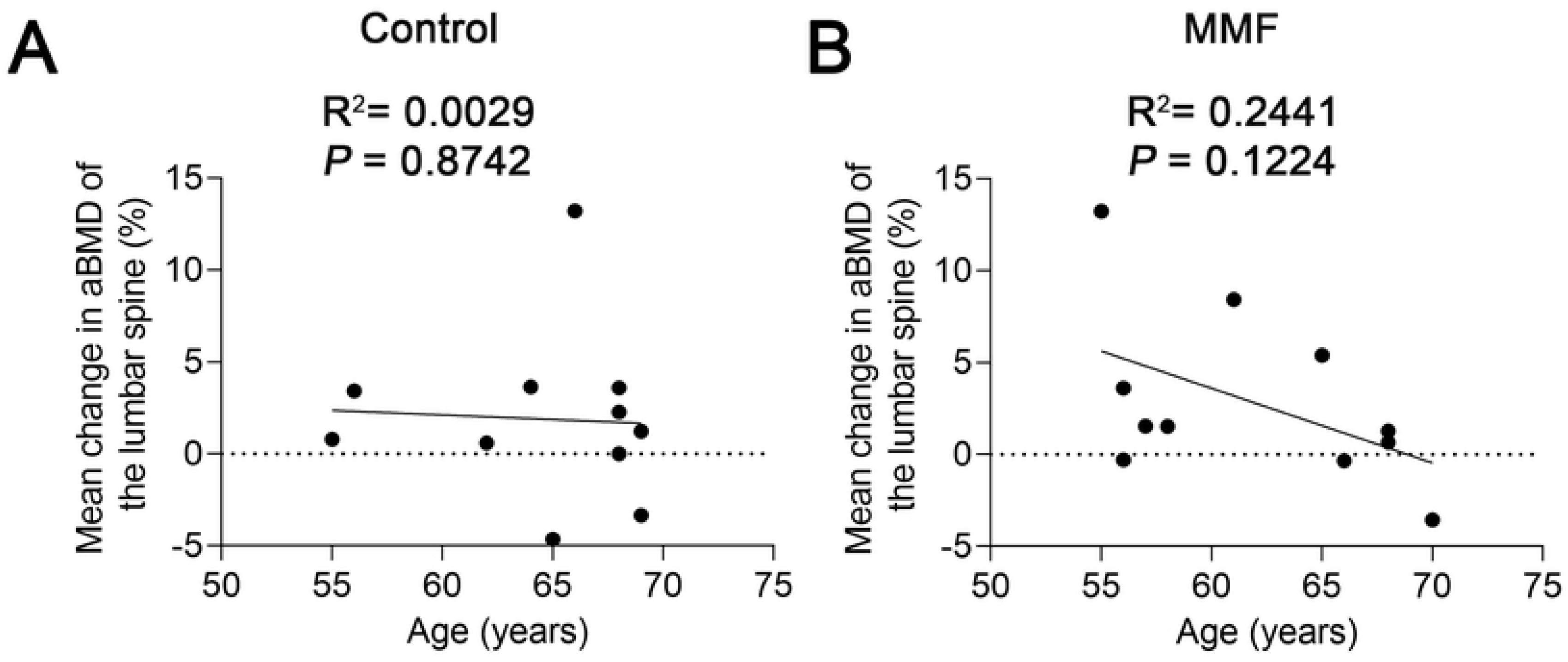
The correlation of participants’ BMD change rate and age after 90 days treatment in clinical trial. (A) Control group. (B) MMF group.

## References

1. Compston, J.E., M.R. McClung, and W.D. Leslie, Osteoporosis. Lancet, 2019. 393(10169): p. 364–376.

2. Song, S., et al., Advances in pathogenesis and therapeutic strategies for osteoporosis. Pharmacol Ther, 2022. 237: p. 108168.

3. van Staa, T.P., et al., Epidemiology of fractures in England and Wales. Bone, 2001. 29(6): p. 517–22.

4. Eastell, R., et al., Postmenopausal osteoporosis. Nat Rev Dis Primers, 2016. 2: p. 16069.

5. Khosla, S. and L.C. Hofbauer, Osteoporosis treatment: recent developments and ongoing challenges. Lancet Diabetes Endocrinol, 2017. 5(11): p. 898–907.

6. Reid, I.R. and E.O. Billington, Drug therapy for osteoporosis in older adults. Lancet, 2022. 399(10329): p. 1080–1092.

7. Cosman, F., et al., Romosozumab Treatment in Postmenopausal Women with Osteoporosis. N Engl J Med, 2016. 375(16): p. 1532–1543.

8. Zhang, W., et al., The Possible Role of Electrical Stimulation in Osteoporosis: A Narrative Review. Medicina (Kaunas), 2023. 59(1).

9. Bassett, C.A., A.A. Pilla, and R.J. Pawluk, A non-operative salvage of surgically-resistant pseudarthroses and non-unions by pulsing electromagnetic fields. A preliminary report. Clin Orthop Relat Res, 1977(124): p. 128–43.

10. Catalano, A., et al., Pulsed electromagnetic fields modulate bone metabolism via RANKL/OPG and Wnt/beta-catenin pathways in women with postmenopausal osteoporosis: A pilot study. Bone, 2018. 116: p. 42–46.

11. Bassett, C.A., R.J. Pawluk, and A.A. Pilla, Acceleration of fracture repair by electromagnetic fields. A surgically noninvasive method. Ann N Y Acad Sci, 1974. 238: p. 242–62.

12. Chen, G., et al., Moderate SMFs attenuate bone loss in mice by promoting directional osteogenic differentiation of BMSCs. Stem Cell Res Ther, 2020. 11(1): p. 487.

13. Degen, I.L. and V.I. Stetsula, Consolidation of bone fragments in a constant magnetic field. Ortop Travmatol Protez, 1971. 32(9): p. 45–8.

14. Wang, S., et al., Safety of exposure to high static magnetic fields (2 T-12 T): a study on mice. Eur Radiol, 2019. 29(11): p. 6029–6037.

15. Dini, L. and L. Abbro, Bioeffects of moderate-intensity static magnetic fields on cell cultures. Micron, 2005. 36(3): p. 195–217.

16. Yang, J., et al., Evidence of the static magnetic field effects on bone-related diseases and bone cells. Prog Biophys Mol Biol, 2023. 177: p. 168–180.

17. Wang, S., et al., Moderate static magnetic field promotes fracture healing and regulates iron metabolism in mice. Biomed Eng Online, 2023. 22(1): p. 107.

18. Aydin, N. and M. Bezer, The effect of an intramedullary implant with a static magnetic field on the healing of the osteotomised rabbit femur. Int Orthop, 2011. 35(1): p. 135–41.

19. Bruce, G.K., C.R. Howlett, and R.L. Huckstep, Effect of a static magnetic field on fracture healing in a rabbit radius. Preliminary results. Clin Orthop Relat Res, 1987(222): p. 300–6.

20. Jiancheng Yang, S.Z., Min Wei, Yanwen Fang, Peng Shang, Moderate Static Magnetic Fields Prevent Bone Architectural Deterioration and Strength Reduction in Ovariectomized Mice. IEEE TRANSACTIONS ON MAGNETICS, 2021. 57(7): p. 9.

21. Scott, J. and E.C. Huskisson, Graphic representation of pain. Pain, 1976. 2(2): p. 175–84.

22. Andreopoulou, P. and R.S. Bockman, Management of postmenopausal osteoporosis. Annu Rev Med, 2015. 66: p. 329–42.

23. Handel, M.N., et al., Fracture risk reduction and safety by osteoporosis treatment compared with placebo or active comparator in postmenopausal women: systematic review, network meta-analysis, and meta-regression analysis of randomised clinical trials. BMJ, 2023. 381: p. e068033.

24. Morley, J., et al., Persistence and compliance with osteoporosis therapies among postmenopausal women in the UK Clinical Practice Research Datalink. Osteoporos Int, 2020. 31(3): p. 533–545.

25. Hiligsmann, M., et al., Determinants, consequences and potential solutions to poor adherence to anti-osteoporosis treatment: results of an expert group meeting organized by the European Society for Clinical and Economic Aspects of Osteoporosis, Osteoarthritis and Musculoskeletal Diseases (ESCEO) and the International Osteoporosis Foundation (IOF). Osteoporos Int, 2019. 30(11): p. 2155–2165.

26. Han, Y., et al., Paracrine and endocrine actions of bone-the functions of secretory proteins from osteoblasts, osteocytes, and osteoclasts. Bone Res, 2018. 6: p. 16.

27. Lacombe, J., G. Karsenty, and M. Ferron, In vivo analysis of the contribution of bone resorption to the control of glucose metabolism in mice. Mol Metab, 2013. 2(4): p. 498–504.

28. Pignolo, R.J., Aging and Bone Metabolism. Compr Physiol, 2023. 13(1): p. 4355–4386.

29. Zhang, B., et al., Biophysical mechanisms underlying the effects of static magnetic fields on biological systems. Prog Biophys Mol Biol, 2023. 177: p. 14–23.

30. Berry M V, G.A.K., Of flying frogs and levitrons. European Journal of Physics, 1997. 18(4): p. 7.

31. Fujimoto, K., et al., The nature of osteoporotic low back pain without acute vertebral fracture: A prospective multicenter study on the analgesic effect of monthly minodronic acid hydrate. J Orthop Sci, 2017. 22(4): p. 613–617.

32. Ohtori, S., et al., Risedronate decreases bone resorption and improves low back pain in postmenopausal osteoporosis patients without vertebral fractures. J Clin Neurosci, 2010. 17(2): p. 209–13.

33. Knezevic, N.N., et al., Low back pain. Lancet, 2021. 398(10294): p. 78–92.

34. Fan, Y., et al., The Analgesic Effects of Static Magnetic Fields. Bioelectromagnetics, 2021. 42(2): p. 115–127.

35. Khoromi, S., et al., Low intensity permanent magnets in the treatment of chronic lumbar radicular pain. J Pain Symptom Manage, 2007. 34(4): p. 434–45.

36. Laszlo, J.F., et al., Effect of local exposure to inhomogeneous static magnetic field on stomatological pain sensation - a double-blind, randomized, placebo-controlled study. Int J Radiat Biol, 2012. 88(5): p. 430–8.

37. Eccles, N.K., A critical review of randomized controlled trials of static magnets for pain relief. J Altern Complement Med, 2005. 11(3): p. 495–509.

38. Dong, D., et al., 16 T high static magnetic field inhibits receptor activator of nuclear factor kappa-Beta ligand-induced osteoclast differentiation by regulating iron metabolism in Raw264.7 cells. J Tissue Eng Regen Med, 2019. 13(12): p. 2181–2190.

39. Yang, J., et al., Regulation of Osteoblast Differentiation and Iron Content in MC3T3-E1 Cells by Static Magnetic Field with Different Intensities. Biol Trace Elem Res, 2018. 184(1): p. 214–225.

40. Zhang, G., et al., 1-2 T static magnetic field combined with Ferumoxytol prevent unloading-induced bone loss by regulating iron metabolism in osteoclastogenesis. J Orthop Translat, 2023. 38: p. 126–140.

41. Lv, H., et al., A static magnetic field improves bone quality and balances the function of bone cells with regulation on iron metabolism and redox status in type 1 diabetes. FASEB J, 2023. 37(7): p. e22985.

